# The role of cholesterol metabolism in predicting clinical outcome of patients with severe COVID-19

**DOI:** 10.1101/2022.11.10.22282185

**Authors:** T.S. Usenko, V.V. Miroshnikova, A.I. Bezrukova, K.S. Basharova, S.B. Landa, Z.R. Korobova, N.E. Liubimova, I.N. Vlasov, M.A. Nikolaev, A.D. Izyumchenko, E.G. Gavrilova, I.V. Shlyk, E.L. Chernitskaya, Y.P. Kovalchuk, P.A. Slominsky, A.A. Totolian, Yu.S. Polushin, S.N. Pchelina

## Abstract

Transcriptomic analysis conducted by us previously revealed upregulation of genes involved in low-density lipoprotein particle receptor (LDLR) activity pathway in lethal COVID-19. Last data suggested the possible role of extracellular vesicles and exomeres in COVID-19 pathogenesis. The aim of the present study was to retrospectively evaluate parameters of cholesterol metabolism as possible predictors of fatal outcome of COVID-19. Blood from 39 patients with severe COVID-19 (the main cohort) were collected at the time of admission to the intensive care unit (ICU) (T1) and 7 days after admission to the ICU (T2). After 30 days patients were divided into two subgroups according to outcome-21 non-survivors and 18 survivors. 28 patients (13 non-survivors and 15 survivors) with severe COVID-19 were included as the replication cohort. The study demonstrated that plasma low-and high-density lipoprotein cholesterol levels (LDL-C and HDL-C) were decreased and CCL20/MIP3ɑ, IL-10, IL-15, IL-27 concentrations were increased in non-survivors compared to controls in T1. *STAB1* gene expression was higher in non-survivors than in survivors (p=0.017) in T2. The conjoint fraction of exomeres and LDL particles measured by dynamic light scattering (DLS) was decreased in non-survivors com-pared to survivors in both the main and replication cohorts. We first showed that change of exomeres fraction may be critical in fatal outcome of COVID-19.

## Introduction

Coronavirus infection (COVID-19) is caused by SARS-CoV-2 virus (severe acute respiratory syndrome coronavirus 2) and significant number of patients with COVID-19 develops critical illness with fatal disease outcome [1]. Despite of the numerous studies devoted to the study of the pathogenesis of COVID-19, there are still many questions, in particular, what factors may determine the lethal outcome. Recent evidence indicates that in the case of SARS-CoV-2 infection immune system can cause a lethal inflammatory response known as cytokine release syndrome or cytokine storm [2]. The intensity of cytokine release determines the clinical severity of the disease, and a pronounced increase in the level of pro-inflammatory cytokines is associated with the development of multiple organ damage to various organs and systems [3]. Last data revealed the link between cholesterol metabolism and susceptibility and severity of COVID-19 [4,5]. In particular, serum lipid levels decrease in patients with COVID-19 and low level of serum low-density lipoprotein cholesterol (LDL-C) have been proposed to be a predictor of poor disease prognosis [4,6]. In our previous study we collected biological samples (plasma and peripheral blood mononuclear cells (PBMCs)) of patients with severe COVID-19 admitted to the intensive care unit (ICU). The inclusion criteria were age between 40 and 80 years with the absence of chronic comorbidities such as cancer, cerebrovascular diseases, heart failure, or renal failure [7]. Transcriptome analysis of PBMCs of patients with COVID-19 upon admission to the ICU revealed a number of differentially expressed genes in patients with different outcomes (survivors and non-survivors). Further to identify key metabolic pathways that may be associated with COVID-19 lethality we used the gene ontology analysis, which demonstrated the activation of the low-density lipoprotein particle receptor (LDLR) activity pathway (GO:0005041) in deceased patients with COVID-19 when compared with survivors [7]. Our results point out this hyperactivation of LDLR activity pathway could be linked to cholesterol metabolism disturbances expressed in changes of lipid spectrum observed in another studies. Additionally, the level of extracellular vesicles (EVs, exosomes) has been estimated [8]. We demonstrated the variety in the size of extracellular vesicles in non-survivors compared to patients with positive outcome, that is in accordance with other studies showing the heterogeneous nature of the exosomes from COVID-19 participants [9,10]. The nomenclature of exosomes remains poorly developed. However, extracellular vesicles are believed to be divided into two subtypes: exosomes with the average size 80-100 nm and also exomeres with a size of less than 50 nm [11]. Exomeres were shown to be very different from exosomes in their lipid composition [12]. Recently we characterized serum exomeres by means of a com-bination of immunoadsorption and dynamic light scattering (DLS) methods [13]. Our results suggested that exomeres play an essential role in cholesterol transport and exchange between the cells. The aim of the current study was to retrospectively evaluate parameters of cholesterol metabolism (plasma lipids concentrations and exomere fraction, the expression levels of LDLR activity pathway genes selected through our previous transcriptome analysis) as well as cytokine profile as possible predictors of fatal outcome in patients with severe COVID-19.

## Materials and Methods

### Approval by institutional research ethics board

This project was approved by Pavlov First Saint-Petersburg State Medical University. A formal written consent form was provided to all included subjects to read and sign prior to the study.

### Subjects

The current main study included 39 patients with severe COVID-19 who were admitted to the to the intensive care unit (ICU) of Pavlov First Saint-Petersburg State Medical University between 1st July 2021 and 31th August 2021. SARS-CoV-2 infection was confirmed through positive RT-PCR in patients. Biological samples (plasma and PBMCs) were collected at the time to admission to the ICU and one week after admission to the ICU. Depending on the disease outcome during the time of observation (30 days) patients were divided into two subgroups: 1) the non-survivor subgroup included 21 deceased patients and 2) the survivor subgroup included 18 patients who survived. The control group included 20 healthy individuals who had never been infected with SARS-CoV-2 that was additionally confirmed by negative PCR and IgG antibody tests. Replication study included 28 patients with severe COVID-19 who were ad-mitted to the ICU of Pavlov First Saint-Petersburg State Medical University from 1st November 2020 to 25th February 2021 and also 20 individuals who were not infected by SARS-CoV-2 at the time of blood collection. Depending on the disease outcome during the time of observation (30 days) patients were divided into two subgroups: 1) the non-survived subgroup included 13 deceased patients and 2) the survived sub-group included 15 patients.

The inclusion criteria were Russian ethnicity, age between 40 and 80 years with the absence of chronic comorbidities such as cancer, cerebrovascular diseases, heart failure, or renal failure but we did not exclude obesity or hypertension, as the common features in patients among patients with severed COVID-19. The characteristics of studies groups are presented in Table 1.

**Table 1.** Characteristics of the studied groups

|  | Main study |  |  |  | Replication study |  |  |  |
| --- | --- | --- | --- | --- | --- | --- | --- | --- |
|  | Non-survivors with COVID-19 (N=21) | Survivors with COVID-19 (N=18) | Controls (N=20) | p<sup>a</sup> | Non-survivors with COVID-19 (N=13) | Survivors with COVID-19 (N=15) | Controls (N=20) | p<sup>a</sup> |
| Mean age ± SE, years | 60.1±9.3 | 55.3±12.9 | 59.3±8.0 | >0.05 | 60.6±10.9 | 57.6±9.1 | 65.2±6.7 | >0.05 |
| Sex (Male:Female) | 10:11 | 11:7 | 11:9 | >0.05 | 10:3 | 9:6 | 12:8 | >0.05 |
| Oxygen saturation * | 81.00±13.61 | 85.89±7.08 | - | >0.05 | 88.1±8.79 | 91.5±4.70 | - | >0.05 |
| T °C * | 37.3±0.86 | 37.5±0.68 | - | >0.05 | 37.0±0.24 | 37.0±0.57 | - | >0.05 |
| CT-SKAN * | 3-4 | 3-4 | - | >0.05 | 3-4 | 3-4 | - | >0.05 |
| Hypertension (yes:no) | 16:5 | 5:13 | - | <0.05 | 7:6 | 8:7 | - | <0.05 |
| Obesity (yes:no) | 11:10 | 2:16 | - | <0.05 | 4:9 | 1:14 | - | <0.05 |
| Neutrophils,<br>×10<sup>9</sup>/l | 6.16<br>(5.31-10.33) | 11.05<br>(6.62-15.36) | - | >0.0<br>5 | 6.26<br>(2.41-12.30) | 9.57<br>(4.03-16.40) | - | >0.0<br>5 |
| Lymphocyte<br>s, ×10<sup>9</sup>/l | 0.60<br>(0.30-0.80) | 0.95<br>(0.60-1.20) | - | <0.0<br>5 | 0.60<br>(0.30-1.40) | 0.85<br>(0.40-1.70) | - | <0.0<br>5 |
| Monocytes,<br>×10<sup>9</sup>/l | 0.29<br>(0.12-0.44) | 0.55<br>(0.28-0.64) | - | >0.0<br>5 | 0.27<br>(0.11-0.91) | 0.42<br>(0.14-1.30) | - | >0.0<br>5 |
| D-dimer,<br>μg/l | 1141<br>(448-2510) | 695 (357-1178) | - | >0.0<br>5 | 1133<br>(273-6692) | 950<br>(287-35200) | - | >0.0<br>5 |
| C-reactive<br>protein, mg/l | 138.5<br>(79.5-202.4) | 142.6<br>(103.4-191.5) | - | >0.0<br>5 | 154 (15-278) | 156 (5-227) | - | >0.0<br>5 |

\* at the time of admission to the ICU, a – analysis of variants (ANOVA), CT-SKAN scores
depending on the extent of consolidation or ground-glass opacities: 1—<25%; 2—25–50%; 3—50–75%; 4—>75%.

### Blood plasma collection

Blood plasma was isolated from peripheral venous blood collected in vacuum containers with EDTA by centrifugation for 20 min, 3000 g. Centrifugation was carried out no later than 20 min after blood taking. Plasma samples were stored at -80°C until the study.

### Plasma cytokine profile

The concentration of cytokines in blood plasma was assessed by multiplex analysis using a MagPix analyzer (Luminex Corporation, USA) using a kit for simultaneous detection of 25 analytes (IL-17F, GM-CSF, IFNγ, IL-10, CCL20/MIP3ɑ, IL-12(p70), IL-13, IL-15, IL-17A, IL-22, IL-9, IL-1β, IL-33, IL-2, IL-21, IL-4, IL-23, IL -5, Il-6, IL-17E/IL-25, IL-27, IL-31, TNFɑ, TNFβ, IL-28A) (Milliplex MAP Human Th17 Magnetic Premix 25 Plex Kit (HT17MG-14K-PX-25, Merk-Millipore, USA) according to the manufacturer’s conditions. The results were processed using xPONENT 4.2 software.

### Plasma lipid profile

Total cholesterol (TC), high-density lipoprotein cholesterol (HDL), low-density lipoprotein cholesterol (LDL), triglycerides (TG) in plasma were determined by OLYMPUS AU400 apparatus.

### Isolation of peripheral blood mononuclear cells

PBMCs were isolated from 8 mL EDTA-anticoagulated venous blood by a Ficoll-Paque gradient method (Ficoll-Paque PLUS, GE Healthcare, Chicago, IL, USA) [14]. After centrifugation, PBMCs were collected from the interface and washed twice with PBS (pH 7.4) to remove the platelet-rich plasma fraction. The PBMCs cell pellets were aliquoted and immediately frozen at −80 °C.

### RNA extraction, cDNA synthesis and quantitative real-time PCR in PBMCs

Total RNA was extracted from PBMCs using GeneJET RNA Purification Kit (K0731, Thermo Fisher Scientific, USA) and complementary DNA (cDNA) was synthesized by Revert Aid First cDNA Synthesis kit (K1622, Thermo Fisher Scientific, USA. Primers and probes were designed by Primer3 program (https://bioinfo.ut.ee/primer3-0.4.0/) (S1 Table). The expression level of cholesterol metabolism genes (*LDLR, PPARG, LRP6, CD36, ANXA2, STAB1*) was assessed with TaqMan probes by real-time quantitative PCR on CFX96 (BioRad, USA). mRNA levels of the studied genes were normalized to mRNA levels of *RPLP0* (Ribosomal Protein Lateral Stalk Subunit P0) and *ACTB* (beta actin). Each sample was tested in triplicate. The value of the expression level relative to the calibrator was determined by the formula: 2-ΔΔC [15].

### Droplet Digital PCR (ddPCR) assay

Nucleic acid copy counts of STAB1 were determined by means of the RPLP0 gene as housekeeping droplets using droplet digital PCR technology (Bio-Rad, USA) following the manufacturer’s protocol and using ddPCR EvaGreen Supermix (Bio-Rad, USA). The thermal cycling conditions for *STAB1* was standard as describe in the manufacturer’s protocol of ddPCR EvaGreen Supermix (Bio-Rad, USA) with annealing temperature 57oC. The ddPCR reaction with primers listed above was performed in a T100 Thermal Cycler. Measurement of positive droplets per µl sample was performed on a QX200 ddPCR Droplet Reader (Bio-Rad, USA). Based on the droplet counts and according to Poisson distribution, absolute nucleic acid copy counts were calculated utilizing the software QuantaSoft (Bio-Rad, USA).

### The method of dynamic light scattering (DLS) for detection of exomeres

Relative levels of exomeres and LDL particles were assessed in plasma by means of a combination of immunoadsorption and dynamic light scattering (DLS) methods as described earlier [13,16]. This approach allows to distinguish particles by size (using hydrodynamic radius) including after preliminary depletion of unnecessary particles by immunoprecipitation with appropriate antibodies to clean particles of interest. Plasma was processed separately with CD9, HSP90 and ApoB-100 anti-bodies with subsequent separation of corresponding particles via centrifugation of 1,000 g for 10 min. Supernatants and initial plasma were analyzed by DLS. Principle of the method is represented in S1 Figure 1. Conjoint fraction of exomeres and LDLs particles is determined by analysis of corresponding peak from measurement of unprocessed plasma (S1 A Figure). Relative level of LDL particles is determined as a mean from analysis of resulting peak after CD9+ and HSP90+ particles (exosomes and exomeres) were removed (S1 B,C Figure). Relative level of exomeres was determined by analysis of resulting peak after ApoB-100 containing particles were removed (S1 D Figure). The measurements were carried out using a laser correlation spectrometer DLS (INTOX MED LLC, St. Petersburg, Russia) with a heterogeneous measurement scheme [17]. Mathematical processing of the obtained data was carried out using the algorithm [17] using the QELSspec (version 3.4) software package, Gatchina, Russia.

**Figure 1.**
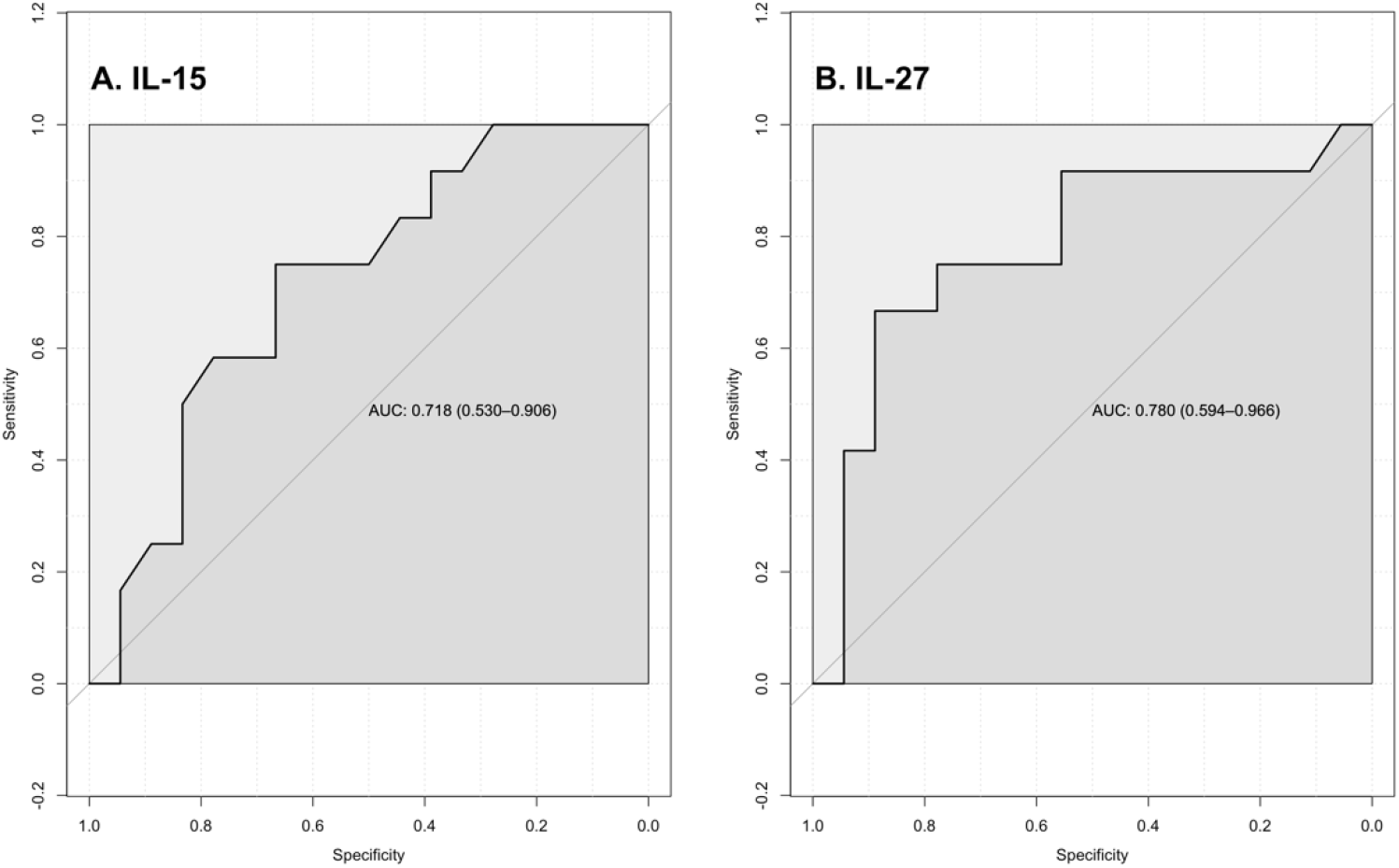
ROC analysis for analytes in blood plasma: A. IL-15, B. IL-27

### Statistical analysis

Conformity of findings to normal distribution was tested using the Shapiro–Wilk test. Comparison of variation between the studied groups was performed by Kruskal-Wallace test. Comparison of variation between two groups was conducted using the nonparametric Mann-Whitney u-test. Correlation analysis was conducted with Spearman coefficient. Significance was established at p<0.05. Statistical analysis was performed using R software (version 3.6.2). Clinical data and experimental data are expressed as the median (min – max).

## Results

This study consists of two independent studies, a main study and a replication study. The main study included the analysis of gene expression profile in PBMC, plasma cytokine profile, plasma lipid profile measured by standard technics and DLS. The replication study included plasma lipid profile measured by standard technics and DLS. All patients admitted to the ICU were characterized with increased secretion of cytokines (cytokine storm), respiratory failure and lung injury with CT-SKAN scores equal to 3-4. The demographic, clinical and laboratory characteristics of the studied populations are shown in Table 1. COVID-19 patients were characterized with lymphopenia and the elevated values of neutrophils, C-reactive protein (CRP) and D-dimer that is in accordance with previous data [18]. There were no differences between survivors and non-survivors except more pronounced lymphopenia in non-survivors in the main study (Table 1).

### Main study

#### Validation of differential gene expression received in transcriptomic data in PBMCs in the studied patient subgroups at the time and 7 days after ad-mission to the ICU and in the control group

We evaluated the expression levels of genes involved in cholesterol metabolism (*PPARG, LDLR, LRP6, CD36, STAB1, ANXA2*) in PBMCs of patients with various outcomes of COVID-19 (convalescence / death), as well as in controls (S2 Table). Studied genes were selected through our previous transcriptomic analysis that revealed upregulation of genes involved in LDLR activity pathway. At the time of admission to the ICU, increased *PPARG* and *CD36* gene expression levels were found both in non-survivors and survivors when compared with controls (*PPARG*: p=0.00024, p=0.00048, respectively; *CD36*: p=0.0027, p<0.0001, respectively), as well as *ANXA2* gene expression level was higher only in survivors (p=0.011) (S5 Table). After 7 days in the ICU increased *STAB1, CD36, LDLR* and simultaneously decreased *ANXA2* gene expression were detected in both non-survivors and survivors compared with the control group (p<0.05). Also, decreased expression levels of *PPARG* and *ANXA2* after 7 days treatment compared with the time of admission were revealed both in non-survivors and survivors (*PPARG*: p=0.016, p=0.016, respectively; *ANXA2*: p=0.00098, p=0.0029, respectively). *LRP6* mRNA level was decreased in survivors when compared with the control group in 7 days after admission to the ICU (p=0.0089). Additionally in survivors *LRP6* decreased 7 days after admission to the ICU when compared with the time to admission the ICU (p=0.02).

It should be noted that only for the *STAB1* gene differential expression between non-survivors and survivors was demonstrated (p = 0.017) and this was specific only for 7 days after admission to the ICU. In non-survivors *STAB1* expression level was increased 7 days after admission to the ICU compared with the time of the admission (p=0.0078). ddPCR was used for confirmation of this result. mRNA copy counts were determined for the *STAB1* gene in non-survived patients with COVID-19, survived patients with COVID-19 in 7 days after admission to the ICU and in control individuals. Increased expression of *STAB1* of non-survivors compared with survivors and controls was revealed (p=0.024, p<0.0001, respectively). So, the result obtained by RT-PCR and demonstrating distinguishing differences between ex-pression level of the *STAB1* gene in non-survivors and survivors was supported by ddPCR (S2 Figure). However, ddPCR did not reveal the differences in expression level of the *STAB1* gene between controls and survived patients with COVID-19 (p>0.05) in 7 days after admission to the ICU.

#### Plasma cytokine profile in the studied patient subgroups at the time to admission to the ICU and in the control group

Plasma cytokine profile was estimated in subgroups of patients with COVID-19 - survivors and non-survivors - at the time of admission to the ICU, as well as the control group. The concentrations of plasma cytokines in the studied groups are presented in Table 2. We found an increased concentration of pro-inflammatory cytokines IL-15, IL-27 in non-survivors when compared with controls and survivors (IL-15: p=0.00014, p=0.00096, respectively; IL-27: p<0.0001, p=0.011, respectively). Secretion of IL-10 and CCL20/MIP3ɑ was increased in non-survivors compared to controls (p=0.0027, p=0.012, respectively). Also, an increase in the concentration of pro-inflammatory cytokines such as IL-15, IL-6, IL-27 was also shown in the subgroup of survived patients with COVID-19 compared with the control group (p=0.049, p=0.026, p=0.00032, respectively). There were no statistically significant differences between the studied groups when comparing the concentration of other analytes in blood plasma (p>0.05).

**Table 2.**
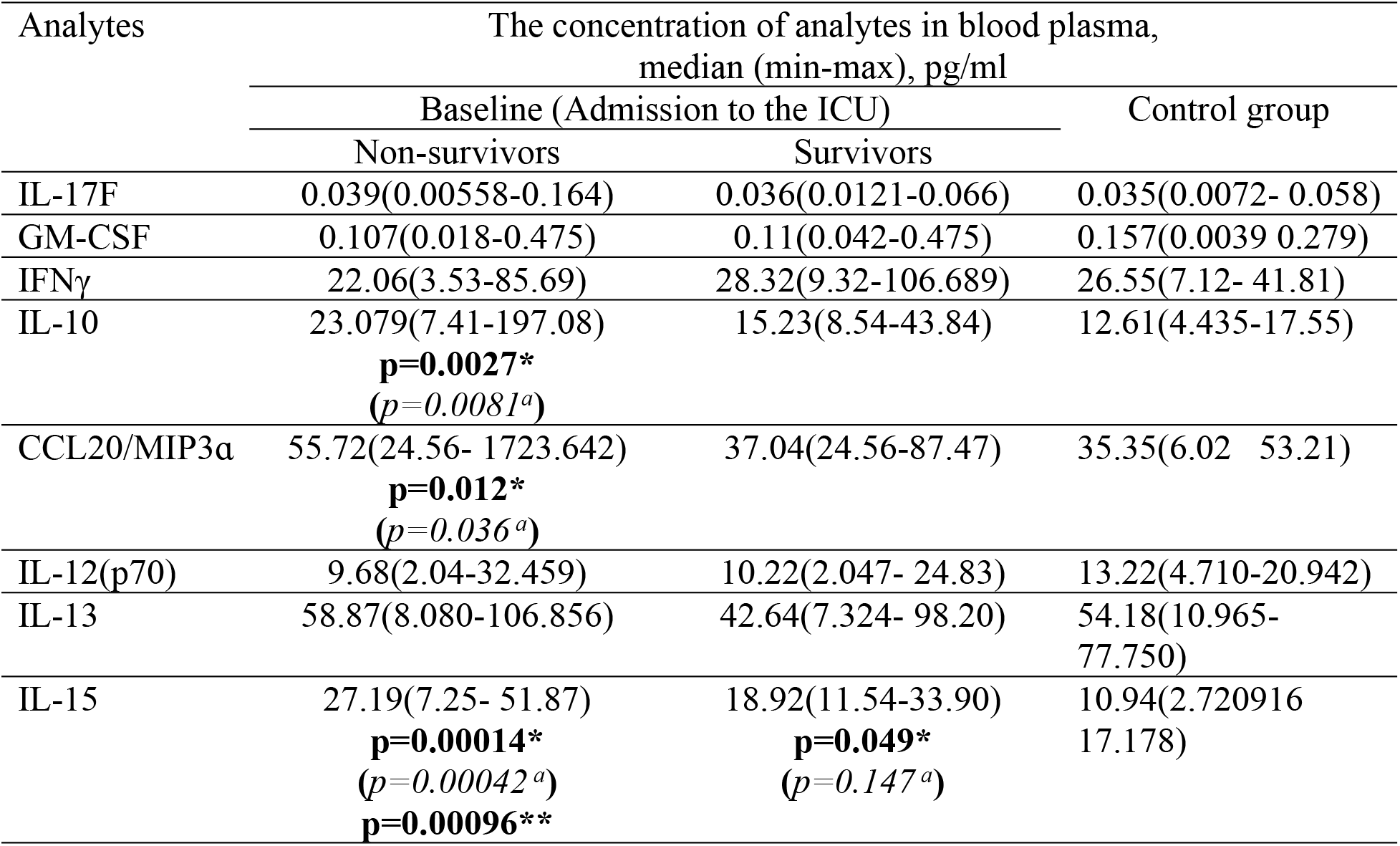
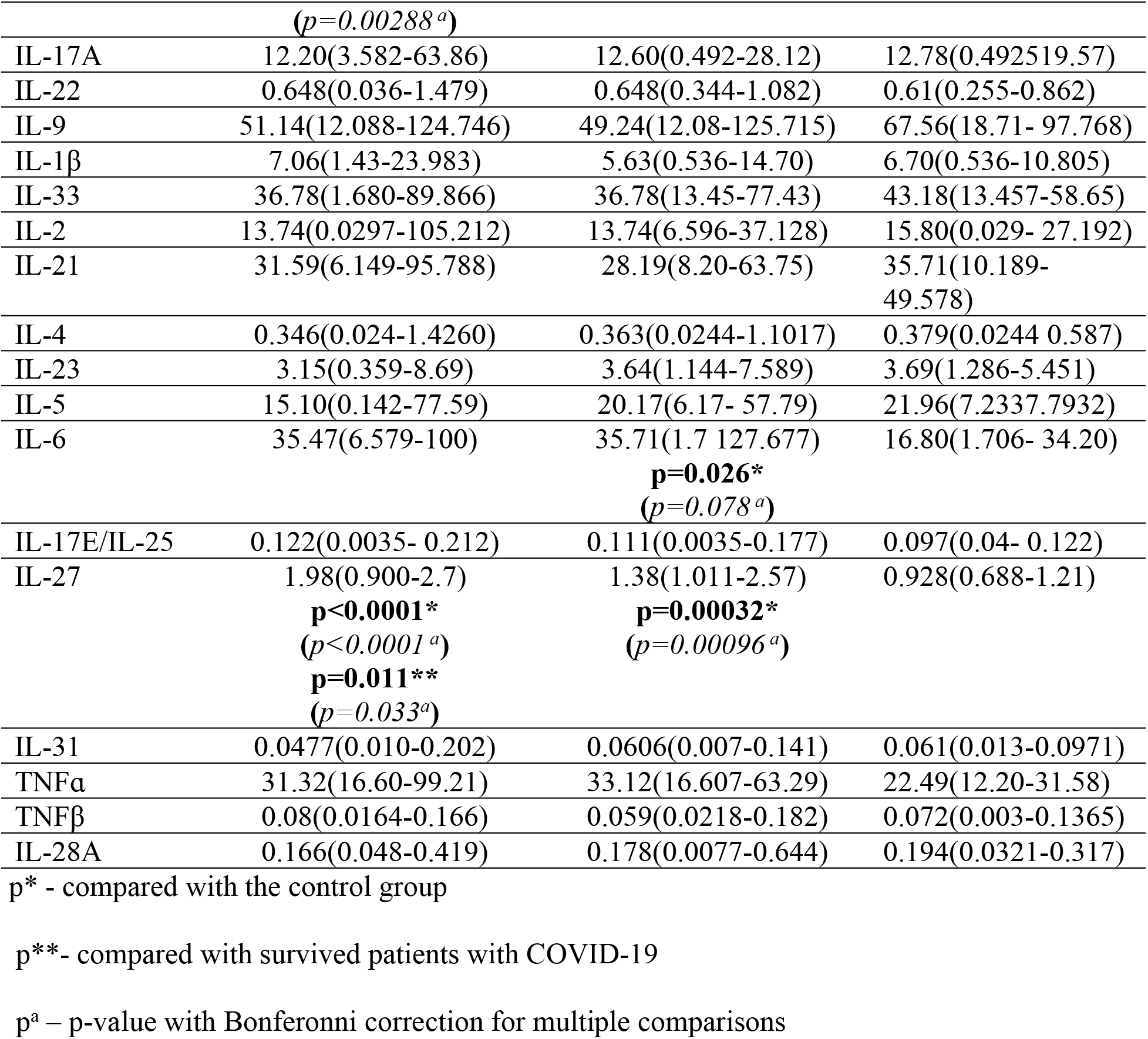
Cytokine profile of blood plasma of the studied groups at the time of admission to the ICU

Thus, non-survivors had higher plasma concentrations of IL-15 and IL-27 than survivors and even higher than controls. So, ROC analysis was performed to determine the threshold value of IL-15 and IL-27 plasma concentrations as predictor of fatal outcome of COVID-19. We have identified a threshold concentration for IL-15 as 25.50 pg/ml. (AUC=0.718 (CI95%:0.53-0.906), specificity =0.75, sensitivity=0.67, accuracy=0.70, p=0.048) (Figure 1 A). The threshold concentration for IL-27 is 1.51 pg/ml (AUC=0.78 (CI95%: 0.594-0.966), specificity=0.67, sensitivity=0.88, accuracy=0.80, p=0.011 (Figure 1 B).

#### Plasma lipid profile in the studied patient subgroups at the time and 7 days after admission to the ICU and in the control group

Plasma lipid profiles of patients with various outcomes of COVID-19 (survivors / non-survivors) at the time of admission as well as in the control group were conducted. The levels of the lipid spectrum are presented in Table 3. We showed a decreased level of LDL and HDL cholesterol in the plasma of non-survived patients with COVID-19 when compared with controls (p=0.0013, p=0.018, p=0.0021, respectively) and tendency to decrease of LDL, HDL and TC levels in survived patients with COVID-19 (p=0.062, p=0.067, p=0.063, respectively). No statistically significant differences in TG concentration between studied groups were found (p>0.05). In order to assess the dynamic change in plasma lipid profile of survivors and non-survivors during severe COVID-19 we additionally evaluated lipid levels in 7 days after admission to the ICU (Table 3). HDL and LDL cholesterol as well as TC levels were decreased in non-survivors compared with survivors (p=0.0043, p=0.0067, p=0.0035 respectively) and the control group. (p=0.001, p=0.0011, p=0.00088, respectively). A pairwise analysis comparing plasma lipid profile change in non-survivors and survivors between two observation points was performed. TC concentration was increased in survivors in 7 days after admission to the ICU when compared with the time of admission (p=0.037). At the same time non-survivors were characterized by continuing decrease of HDL and LDL cholesterol levels in 7 days after admission to the ICU when compared with the time of admission (p=0.049, p=0.0014, respectively). Plasma TG concentration did not differ between the studied groups neither at the time of admission nor 7 days after (p>0.05).

**Table 3.** Plasma lipid profiles in the studied groups

| Parameter<br>s | Lipid spectrum of blood plasma,<br>median (min-max), mmol/l |  |  |  | Control<br>group |
| --- | --- | --- | --- | --- | --- |
|  | Admission to the ICU |  | 7 days after admission to the<br>ICU |  |  |
|  | Non-survivors | Survivors | Non-survivors | Survivors |  |
| TC | 3.50(2.10-5.79)<br>**p=0.0063\*** | 4.19(2.50-6.70) | 2.80(1.5-5.10)<br>**p=0.00088\***<br>**p=0.0035\*\*** | 5.00(3.9-7.6)<br>**p=0.037\*\*\*\*** | 5.21(2.50-6.59) |
| LDL | 1.81(0.14-3.52)<br>**p=0.0013\*** | 2.22(0.92-4.32)<br>**p=0.062\*** | 1.36(0.21-1.18)<br>**p=0.0011\***<br>**p=0.0043\*\***<br>**p=0.0014\*\*\*** | 2.68(1.80-4.681) | 3.24(0.71-4.87) |
| HDL | 0.83(0.27-1.39)<br>**p=0.018\*** | 0.89(0.56-1.46)<br>**p=0.067\*** | 0.63(0.21-1.18)<br>**p=0.001\***<br>**p=0.0067\*\***<br>**p=0.049\*\*\*** | 1.06(0.61-1.30) | 1.09(0.68-1.95) |
| TG | 2.09(0.88-4.28) | 1.66(0.89-4.50) | 2.62(1.41-3.40) | 2.94(1.02-9.66) | 1.59(0.93-2.78) |

\* - compared with controls, \*\* - compared with survivors with COVID-19 (in a week), \*\*\* - compared with non-survivors with COVID-19 (admission), \*\*\*\* - compared with survivors with COVID-19 (admission)

#### Relative levels of plasma exomeres and LDL particles detected by means of DLS in the studied patient subgroups at the time and 7 days after admission to the ICU and in the control group

Exomeres are a class of extracellular vesicles enriched in cholesterol and comparable in size with LDLs. Our approach allowed to distinguish particles by size (using hydrodynamic radius). Relative levels of exomeres and LDLs as well as conjoint fraction of exomeres and LDL particles were assessed by alternative method combination of immunoadsorption and DLS. We analyzed conjoint level of particles with hydrodynamic radius of 20 nm (exomers and LDLs), next we analyzed the same peak after immunoadsorption of exomeres using antibodies against CD9 and HSP90 (LDL particles remain after cleaning) and antibodies against apolipoproteins B100 (exomeres remain after cleaning). It should be noted that relative LDL level determined by this method as relatively real quantity of particles is highly positively correlated with those measured by standard technique. Level of plasma exomeres was decreased in non-survivors compared with survivors at time to admission to the ICU (p=0.01), still there were not any differences between the studied groups in 7 days after admission (Figure 2, S3 Table). Level of LDL particles was decreased in non-survivors at the time of admission to the UCI compared with controls (p=0.01). 7 days after to admission to the ICU level of LDL particles was lower in non-survivors compared with survivors (p=0.005) and controls (p=0.004). Interestingly, if one takes into account conjoint fraction of exomeres and LDL particles (ExoM_LDL) significant reduction of this parameter in non-survivors when compared with survivors and the control group is traced both at the time of admission to the ICU (p<0.0001, p<0.0001, respectively) and 7 days after (p=0.00043, p=0.0089, respectively).

**Figure 2.**
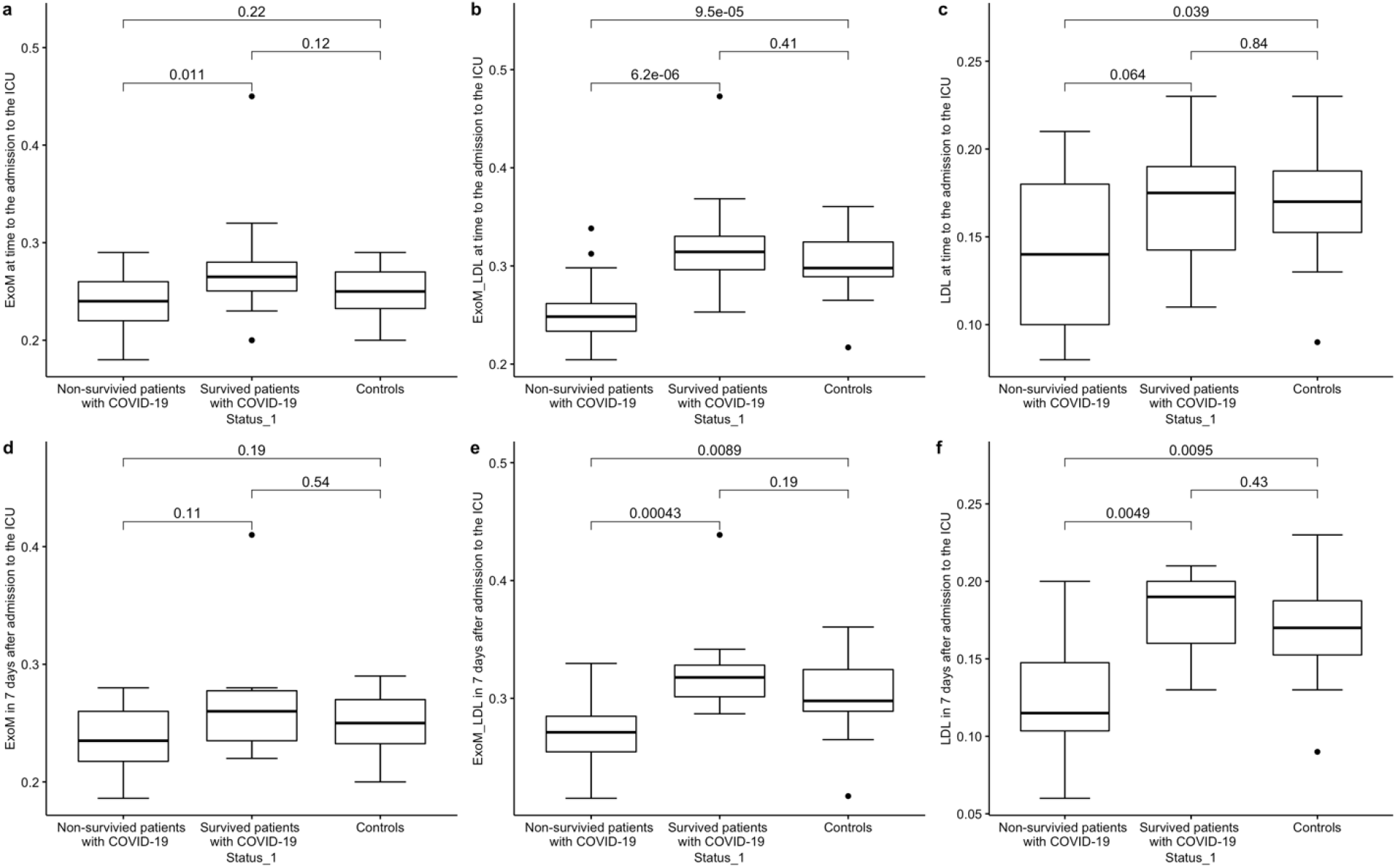
Relative level of plasma exomeres with concentration of TC bound by LDL: a. ExoM (admission to the ICU), b. ExoM_LDL (admission to the ICU), c. LDL (admission to the ICU), d. ExoM (7 days after admission to the ICU), e. ExoM_LDL (7 days after admission to the ICU), f. LDL (7 days after admission to the ICU)

#### Survival Predictive Value Analysis of the parameters of lipid profile measured by DLS and standard technic by ROC analysis

ROC analysis was conducted to determine the threshold value of lipid concentrations (TC, LDL, HDL) measured by standard technique between survived and non-survived patients. Significant threshold value was not found (data are not presented). Also, ROC was performed to determine the threshold value of relative level of exomeres, LDL particles and conjoint fraction of exomeres and LDLs measured by DLS. Among all studied parameters reflecting lipid spectrum conjoint fraction of exomeres and LDL particles was the best to distinguish non-survivors and survivors at two estimated points (the time and 7 days after to admission to the ICU) (Figure 3). We have identified a threshold value for ExoM_LDL at time to admission to the ICU as 0.272 (AUC=0.894 (CI 95%:0.792-0.997), specificity =0.809, sensitivity=0.889, accuracy=0.846, p= 6.242773e-06) (Figure 3 A). The threshold value for ExoM_LDL 7 days after the time to admission to the ICU was 0.284 (AUC=0.917 (CI 95%: 0.792-1.000), specificity=0.75, sensitivity=1.00, accuracy=0.80, p= 0.00043) (Figure 3 B). Using the logistic regression model to predict the probability of negative outcome for three parameters, IL-15, IL-27 and conjoint fraction of exomeres and LDL at the time of admission to the ICU, AUC was calculated as 0.958 (S3 Figure). The combination of IL-15, IL-27 concentrations and conjoint fraction of exomeres and LDL measured at the time to admission to the ICU was shown as the most sensitive marker of lethal outcome of COVID-19.

**Figure 3.**
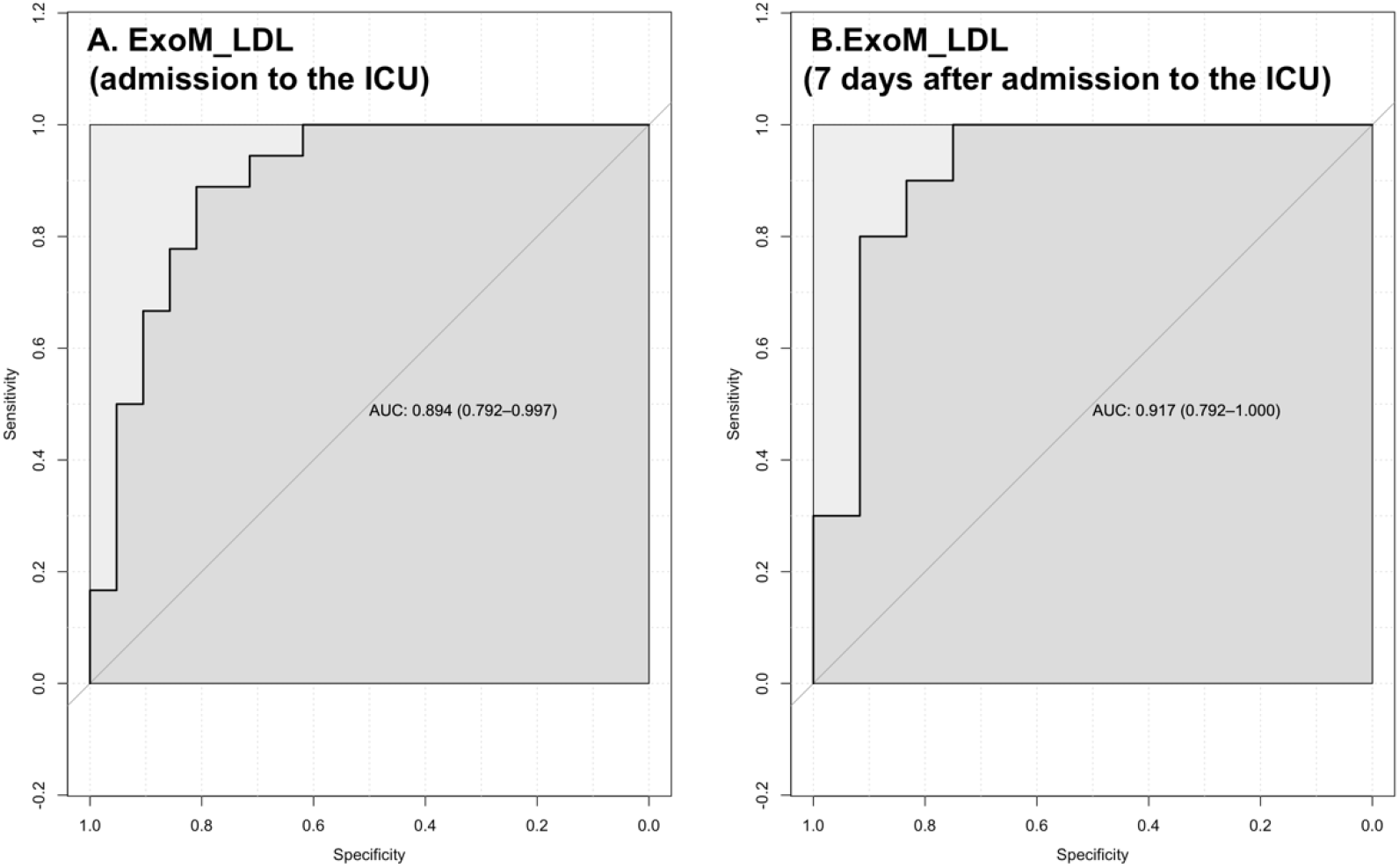
ROC analysis for analytes in blood plasma: A. ExoM_LDL (admission to the ICU), B. ExoM_LDL (7 days after admission to the ICU).

#### Correlation analysis between lipid profile measured by DLS and standard technic

The matrices of correlation pairs of lipid profile and the ratio of the contributions to scattering (SIC) parameters for exomeres and LDL particles (ExoM, ExoM_LDL, LDL) in the three above-mentioned groups (non-survivors with COVID-19, survivors with COVID-19, controls) in both time points are presented in S4 Table and S5 Table. We found a strong positive correlation between LDL measured by DLS and standard technique, namely, between SIC LDL (rplU) with plasma LDL in all studied groups in both time points (admission to the ICU, 7 days after to the ICU) (S4 Table, S5 Table).

#### Analysis of correlation between plasma lipid parameters and cytokine/chemokine concentrations

Interleukins 6, 10, 15, 27 and chemokine CCL20/MIP3ɑ which plasma concentrations were significantly increased in patients with COVID-19 compared with controls were selected for correlation analysis with parameters of lipid profile and also conjoint fraction of exomeres and LDL particles. Spearman’s correlation analysis was conducted for all groups. Mostly negative correlation between selected inflammatory markers and lipid concentrations were observed in non-survivors (S4 Figure). HDL correlated negatively with IL-15 (r=-0.495, p=0.036), IL-27 (r=-0.692, p=0.0015) and LDL correlated negatively with IL-15 (r=-0.278, p=0.026) and LDL-rlpU with IL-27 (r=-0618, p=0.006) in non-survivors with COVID-19 at time of admission to the ICU. Negative correlation was found in group of non-survivors 7 days after admission to the ICU between TC, HDL, LDL and IL-15 (r=-0.790, p=0.011; r=-0.702, p=0.032; r=-0.755, p=0.019, respectively) and IL-27 (r=-0.882, p=0.002; r=-876, p=0.002; r=-0.948, p=0.00009, respectively). Also, LDL-rlpU negative correlated with IL-27 (r=-0.794, p=0.006), conjoint fraction of exomeres and LDL (ExoM_LDL) with CCL20/MIP3ɑ (r=-0.661, p=0.0373).

### Replication study

#### Plasma lipid profile in the studied patient subgroups at the time and 7 days after admission to the ICU and in the control group

Plasma lipid profiles of patients with various outcomes of COVID-19 (survivors / non-survivors) at the time of admission as well as in the control group from replication study are presented in Table 4.

**Table 4.**
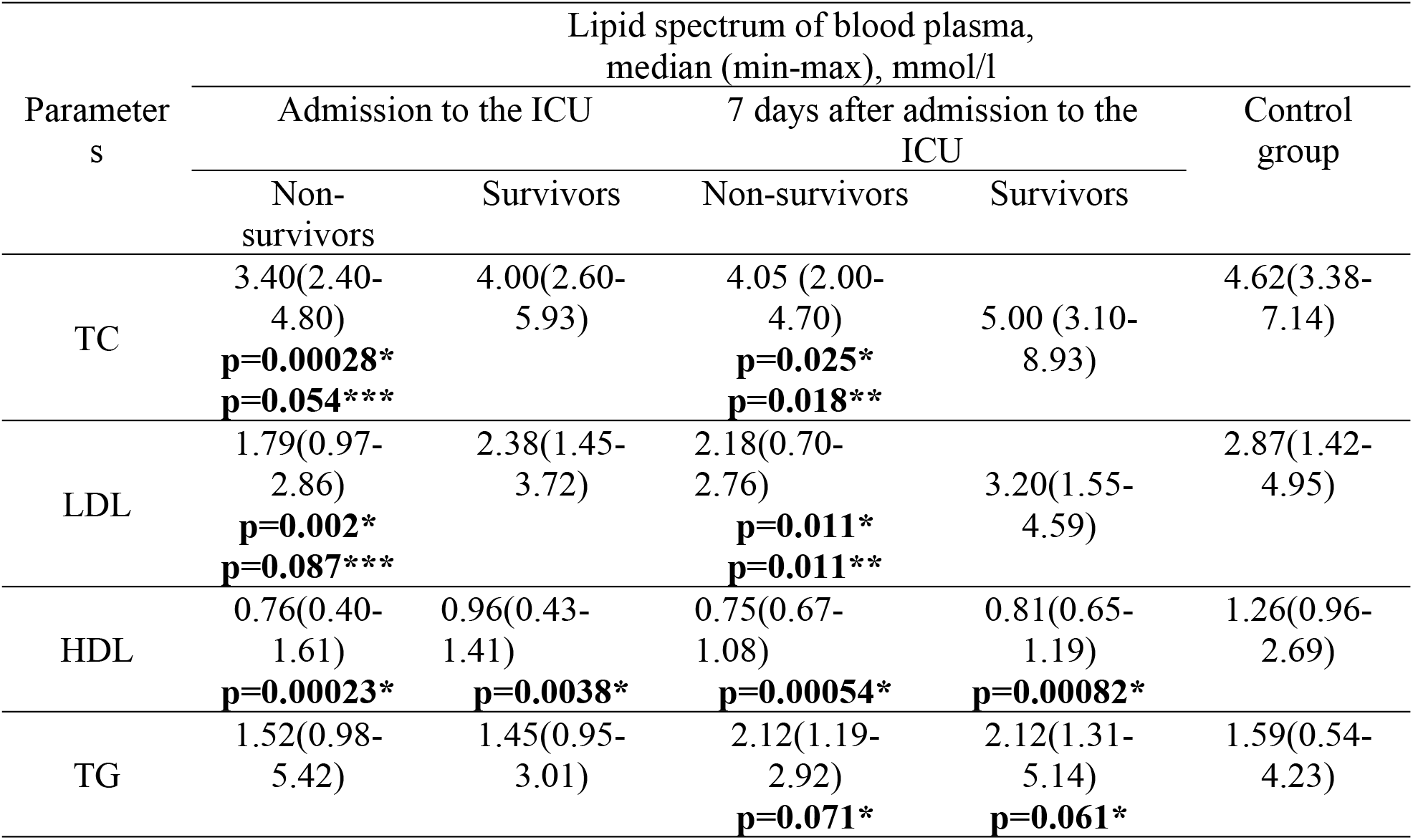
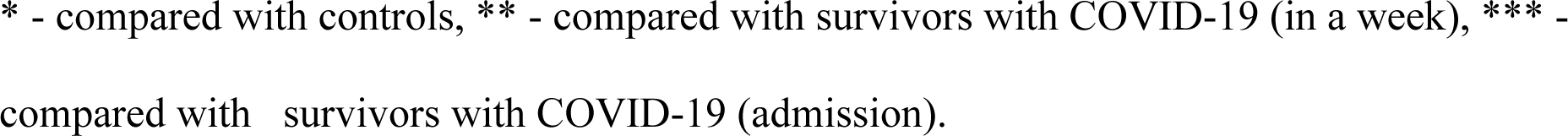
Plasma lipid profiles in the studied groups

| Parameter<br>s | Lipid spectrum of blood plasma,<br>median (min-max), mmol/l |  |  |  | Control<br>group |
| --- | --- | --- | --- | --- | --- |
|  | Admission to the ICU |  | 7 days after admission to the<br>ICU |  |  |
|  | Non-survivors | Survivors | Non-survivors | Survivors |  |
| TC | 3.40(2.40-4.80)<br>**p=0.00028\***<br>**p=0.054\*\*\*** | 4.00(2.60-5.93) | 4.05 (2.00-4.70)<br>**p=0.025\***<br>**p=0.018\*\*** | 5.00 (3.10-8.93) | 4.62(3.38-7.14) |
| LDL | 1.79(0.97-2.86)<br>**p=0.002\***<br>**p=0.087\*\*\*** | 2.38(1.45-3.72) | 2.18(0.70-2.76)<br>**p=0.011\***<br>**p=0.011\*\*** | 3.20(1.55-4.59) | 2.87(1.42-4.95) |
| HDL | 0.76(0.40-1.61)<br>**p=0.00023\*** | 0.96(0.43-1.41)<br>**p=0.0038\*** | 0.75(0.67-1.08)<br>**p=0.00054\*** | 0.81(0.65-1.19)<br>**p=0.00082\*** | 1.26(0.96-2.69) |
| TG | 1.52(0.98-5.42) | 1.45(0.95-3.01) | 2.12(1.19-2.92)<br>**p=0.071\*** | 2.12(1.31-5.14)<br>**p=0.061\*** | 1.59(0.54-4.23) |

\* - compared with controls, \*\* - compared with survivors with COVID-19 (in a week), \*\*\* - compared with survivors with COVID-19 (admission).

TC, LDL and HDL cholesterol in the plasma were decreased in non-survived patients with COVID-19 compared to controls (p=0.00028, p=0.002, p=0.00023. respectively) and TC and LDL had a tendency to decrease compared with survived patients with COVID-19 (p=0.054, p=0.087, respectively) at the time to admission to the ICU. Also, we demonstrated a decrease of plasma HDL cholesterol in survived patients with COVID-19 when compared with controls (p=0.0038). No statistically significant differences in TG concentration between studied groups were found (p>0.05).

In 7 days after admission to the ICU TC and LDL concentration were decreased in non-survivors compared with controls (p=0.025, 0.011, respectively) and survivors (p=0.018, p=0.011, respectively). HDL concentration was decreased and TG concentration had tendency to increase in non-survivors and survivors compared with controls (p=0.00054, p=0.071; p=0.00082, p=0.061, respectively).

A pairwise analysis comparing plasma lipid profile change in non-survivors and survivors between two observation points (the time and 7 days after admission to the ICU) did not reveal significant differences(p>0.05).

#### Relative levels of plasma exomeres and LDL particles detected by means of DLS in the studied patient subgroups at the time and 7 days after admission to the ICU and in the control group

Level of plasma exomeres (ExoM) was decreased in non-survivors compared with survivors and controls at time to admission to the ICU (p=0.0037, p=0.0089, respectively), still the significant differences between non-survivors and controls were persisted with control group in 7 days after admission (p=0.031) (Figure 4, S6 Table). Level of LDL particles was decreased in non-survivors and survivors at time to admission to the UCI compared with controls (p=0.0026, p=0.019, respectively). 7 days after to admission to the ICU the level of LDL particles was lower in non-survivors compared with survivors (p=0.0037) and controls (p=0.021). Conjoint fraction of exomeres and LDL particles (ExoM_LDL) was significantly reduced in non-survivors when compared with survivors and the control group (p=0.011, p=0.00039, respectively) and in survivors compared with controls (p=0.025) at time to admission to the ICU. 7 days after admission to the ICU ExoM_LDL was decreased in non-survivors but not in survivors compared to controls (p=0.012).

**Figure 4.**
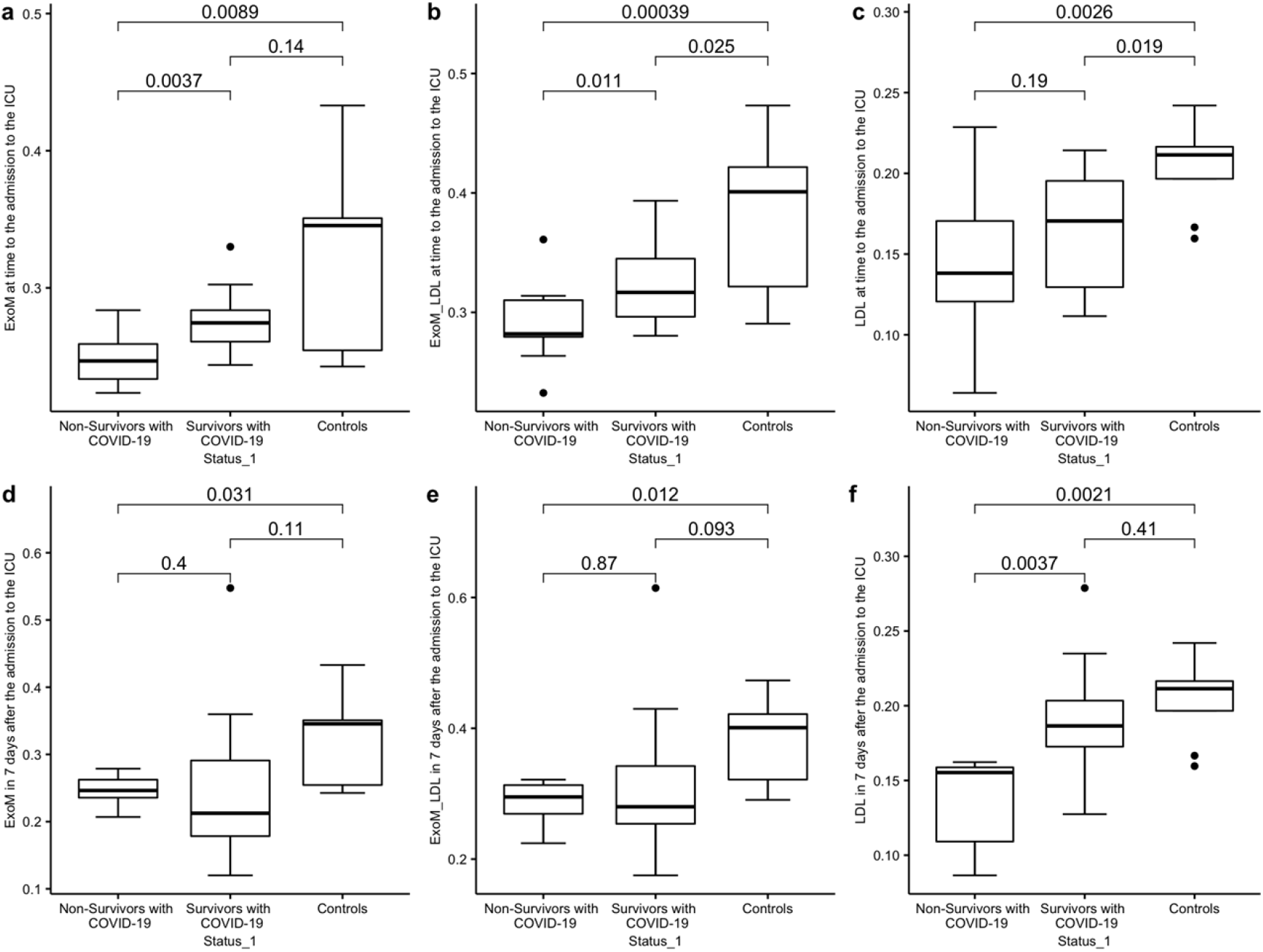
Relative level of plasma exomeres with concentration of TC bound by LDL for replication study: a.ExoM (admission to the ICU), b. ExoM_LDL (admission to the ICU), c. LDL (admission to the ICU), d. ExoM (7 days after admission to the ICU), e. ExoM_LDL (7 days after admission to the ICU), f. LDL (7 days after admission to the ICU).

#### Survival predictive value of conjoint fraction of exomeres and LDLs measured by DLS using ROC analysis

ROC analysis for replication study was performed to determine the threshold value of relative level of conjoint fraction of exomeres and LDLs (Figure 5). We have identified a threshold value for ExoM_LDL at time to admission to the ICU as 0.296 (AUC=0.762 (CI 95%:0.575-0.948), specificity =0.692, sensitivity=0.800, accuracy=0.750, p=0.0199) (Figure 5).

**Figure 5.**
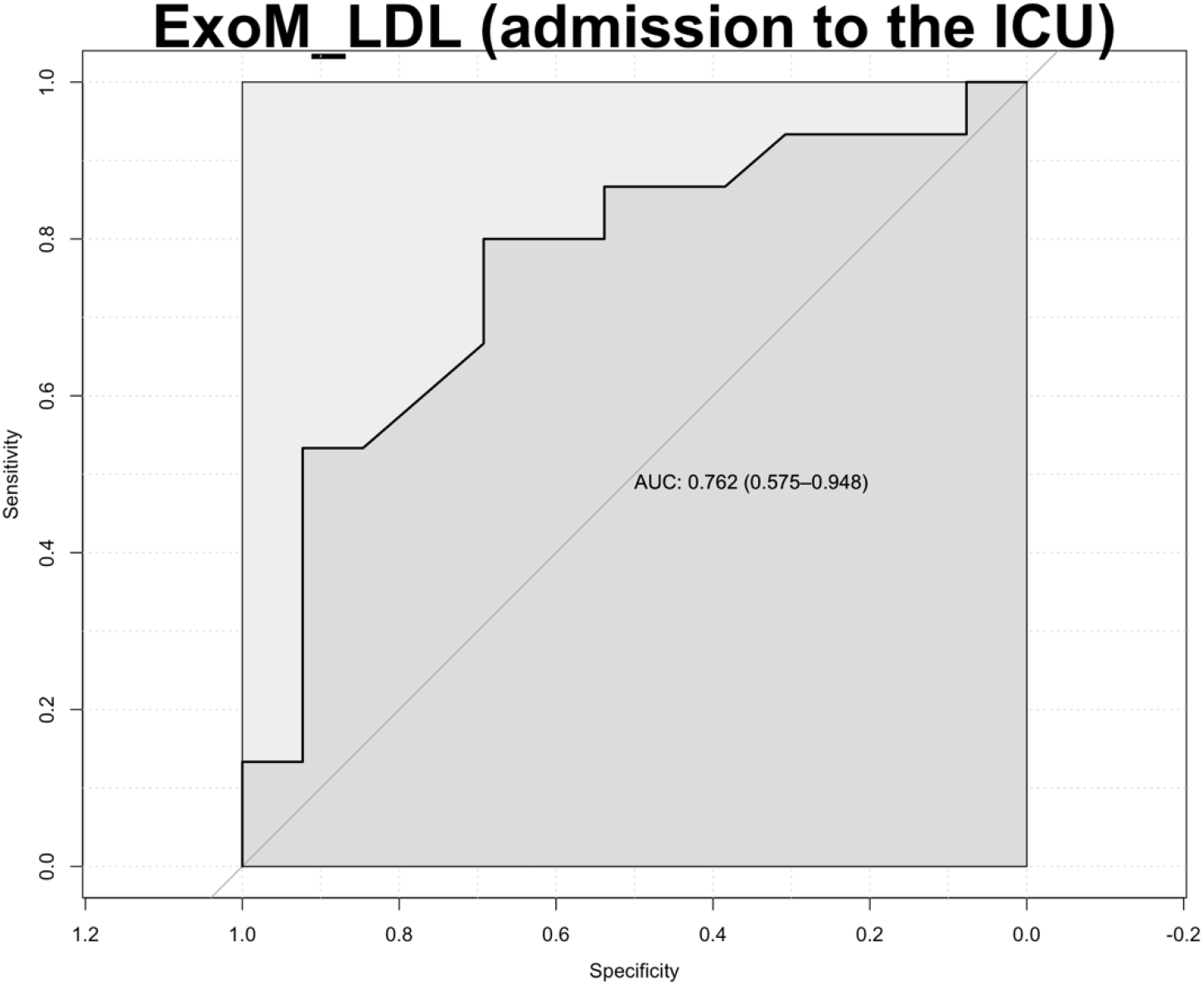
ROC analysis for ExoM_LDL in blood plasma (admission to the ICU)

#### Correlation analysis between lipid profile measured by DLS and standard technics

The matrices of correlation pairs of lipid profile and the ratio of the contributions to scattering (SIC) parameters for exomeres (ExoM, ExoM_LDL, LDL) in the three above-mentioned groups (non-survivors with COVID-19, survivors with COVID-19, controls) in both time points are presented in S7 Table and S8 Table. We revealed a positive correlation between LDL measured by DLS and standard technique, namely, between SIC LDL (rplU) with plasma LDL in non-survivors and survivors at time to admission to the ICU and in survivors 7 days after admission to the ICU (S7 Table, S8 Table). Interesting, conjoint fraction of exomeres and LDL particles (ExoM_LDL) was strong positive correlated with TC in both time points (admission to the ICU, 7 days after to the ICU) in all studied groups and with LDL at time to admission to the ICU in all studied groups and in non-survivors 7 days after admission to the ICU (S7 Table, S8 Table).

#### Prediction of fatal outcome of COVID-19 using multivariable regression analysis (main study, replication study)

Multivariable regression analysis was performed to identify contribution of all studied parameters (cytokines profile, lipid profile, gene expression profile, exomeres) to fatal outcome of COVID-19 in patients, adjusted for hypertension, obesity, age and sex (S9 Table). Relative level of conjoint fraction of exomeres and LDL particles (ExoM_LDL) was strong associated with fatal outcome of COVID-19 (OR=2.22e+04, 95%CI: 1.82-2.69e+08; p=0.0395) at time to admission to the ICU in full regression model in the main study (S9 Table).

Also, we estimated contribution of the parameters of lipid profile and exomeres to fatal outcome of COVID-19 in patients, adjusted for hypertension, obesity, age and sex in the main study and the replication study (S10 Table and S11 Table). We found the association between the higher relative level of conjoint fraction of exomeres and LDL particles (ExoM_LDL) and lower risk of fatal outcome of COVID-19 at two observation points (admission to the ICU, 7 days after to admission to the ICU) in the main study (OR=1.075, CI95%:1.032-1.120, p=0.00146; OR=1.089, CI95%:1.022-1.159, p=0.017, respectively) and at time to admission to the ICU in the replication study (OR=577.779, CI95%:4.471-74621.06, p=0.0185) that is in accordance with our results described in the paragraphs 3.1.4 and 3.2.2.

## Discussion

Here, we for the first time carried out simultaneous assessment of the complex of parameters that determine cholesterol metabolism in order to find relevant biomarkers of poor prognosis and fatal outcome of severe COVID-19. At first, we conducted verification of our previous genomic data on gene expression in patients survived or deceased after COVID-19 by means of two approaches, real-time PCR and ddPCR, and additionally estimated the dynamics of lipid profile with standard enzymatic assay along with alternative parameters - relative levels of plasma exomeres as well as conjoint fraction of exomeres and LDL particles measured by DLS in patients with severe COVID-19 during the stay in the ICU. Plasma cytokine profile (25 analytes) was detected by means of Luminex assay upon admission.

Our previous transcriptomic data allowed us to select gene set associated with poor prognosis of COVID-19 and subsequently in this study *PPARG, LDLR, LRP6, CD36, STAB1, ANXA2* gene expression was shown to be changed in different time points analyzed.

The most interesting finding of this work is that increased expression levels of LDL receptors genes (*LDLR, CD36, STAB1*) were found in non-survivors compared to controls. *CD36* gene expression was increased in non-survivors as well as survivors at the admission to the ICU, and remained elevated in 7 days. *LDLR* gene expression was markedly increased in 7 days in the ICU both in survivors and non-survivors. As for the *STAB1* gene its expression was increased only in 7 days in the ICU but at the same time distinguished non-survivors from survivors. While *LDLR* encodes canonical LDL receptor *CD36* and *STAB1* encode important scavenger receptors for modified, primarily oxidized LDL (oxLDL). It was shown earlier that COVID-19 severity is associated with oxidative stress and products of lipid peroxidation accumulate during disease [19]. Thus, elevated CD36 and STAB1 gene expression may indicate increased oxidized lipoproteins uptake during severe COVID-19 and this in turn partially could explain lipid profile abnormalities ob-served in our study as well as in others. In particular plasma TC, LDL and HDL cholesterol levels were markedly reduced in non-survivors when compared with the control group at the time of admission to the ICU and 7 days after. And while lipid concentrations tended to go up in survivors during treatment in the ICU they were continuously reduced in non-survivors.

*STAB1* gene expression after 7 days after admission to ICU was associated with fatal outcome of COVID-19. Survived patients also have increased *STAB1* gene expression compared with controls but significantly lower than in non-survivors that supported the results of our previous transcriptome analysis [7]. Stabilin 1 encoded by *STAB1* is a multifunctional transmembrane receptor and functions simultaneously as class H scavenger receptor and an adhesion molecule. So, stabilin 1 actively participate in lipid metabolism mediating oxLDL clearance but also play important role in inflammation [20]. Stabilin-1 is involved in regulating T and B lymphocyte movement to sites of inflammation. Its expression may be inducible in reaction to diverse proinflammatory stimuli linked of course of COVID-19 infection and observed hyperexpression in non-survivors may be a consequence of cytokine storm.

Downregulation of the annexin A2 (*ANXA2*) gene expression in all patients with severe COVID-19 7 days after stay in the ICU in spite of its moderate baseline in-crease only in survived patients was also demonstrated in this study. Annexin A2 has been implicated in the inflammatory process by playing a crucial role in the clearance of apoptotic lymphocytes [21,22]. Annexin A2 modulates phospholipid membrane composition in particular participating in the recruitment of cholesterol and glyco-sphingolipids into larger clusters [23]. Thus, its downregulation in 7 days of time point may reflect lipid depletion state in severe COVID-19.

*PPARG* gene upregulation also could be one of the factors responsible for this COVID-19-specific lipid profile linking it to the inflammation. PPARγ has been involved in lipid metabolism and inflammatory responses [24]. PPARγ belongs to the nuclear receptor family of ligand-activated transcription factors and is highly ex-pressed in the immune cells [25]. Immunomodulatory function of PPARγ includes participating in resolution of inflammation so its activation was supposed to be a sign of inflammation in the lipid depleted setting that is characteristic of severe COVID-19 [26]. It should be noted PPARG gene expression was upregulated in survivors and non-survivors compared with the control group at the time of admission but flattened after 7 days in the ICU and could be related to the course of the disease. PPARγ directly regulates the expression of genes involved in lipid transport and metabolism including CD36 [27]. Observed increased CD36 gene expression in both survivors and non-survivors may be a consequence of PPARγ activation and here with remains increased after 7 days in the ICU. By targeting inflammation, ACE2 and the WNT/β-catenin pathway, PPARγ agonists were suggested to be interesting candi-dates for SARS-CoV-2 therapy due to modulation of the cytokine storm by preventing the cytokine overproduction [28,29].

It is known that SARS-CoV-2 virus particles maturing in infected alveolar ep-ithelial cells through exocytosis enter the lung tissue, which leads to the release of a large number of pro-inflammatory cytokines (cytokine storm), infiltration of the lung tissue by macrophages, neutrophils and T cells and development acute respiratory distress syndrome, which is accompanied by severe impairment of lung function with a decrease in blood oxygen saturation [1]. The severity of the cytokine storm de-termines the clinical severity of the disease. Our study also found increased secretion of pro-inflammatory cytokines such as CCL20/MIP3ɑ, IL-10, IL-15, IL-27 in non-survivors than in controls. Increased secretion of CCL20/MIP3ɑ is a new finding in COVID-19, although we previously reported an increase in concentrations of other macrophage inflammatory proteins, CCL3/MIP-1a and CCL4/MIP-1b [30]. CCL20/MIP3ɑ mainly elicits its effector function via CCR6 receptor binding; interestingly enough, we previously noted a negative correlation between CCR6 ex-pressing CD8+ cells and IL-27 [31]. However, secretion of IL-15 and IL-27 was increased in non-survivors compared to both survived patients and controls that is consistent with previously published data [32]. It is interesting to note that IL-15 and IL-27 are early cytokines, and this change in their concentration indicates the activation of the innate immune response when the virus is recognized by the epithelial or dendritic cells of the body, followed by the induction of the innate defense of body and induction of inflammation processes [31,33–34]. In accordance with our and previous data, IL-15, IL-27 were previously associated with the severity and fatal outcome of COVID-19 [32, 35–38]. Additionally, we revealed the negative correlation between concentration of these cytokines and key lipid parameters (TC, LDL, HDL), in particular, LDL cholesterol level, and this correlation was more pronounced after 7 days in the patient subgroup with fatal outcome. It is hypothesized that pro-inflammatory cytokines induced by viral infection may modulate lipid metabolism, including LDL oxidation by releasing reactive oxygen species to facilitate LDL clearance by cells [39]. On the other hand, an inflammatory response could be a consequence of the lipid alterations. Increased PBMC expression levels of *LDLR, CD36, STAB1* observed in non-survivors in our study could promote the increase of lipid uptake and subsequent formation of lipid droplets. At the same time lipid droplets were shown to be involved in RNA viruses’ replicative cycle, in inflammatory mediator production and innate signaling in immune cells that could explain link between cytokine overproduction and plasma lipid profile alteration [40]. Monocytes from COVID-19 patients showed an increased lipid droplet accumulation compared to SARS-CoV-2 negative donors [40]. Dysregulated monocyte responses play a pivotal role in the uncontrolled cytokine/chemokine production observed during severe COVID-19 [40]. Still our results were based on a single Russian cohort of COVID-19 patients; future studies will be needed for extending the understanding of interplay of lipid dysregulation and cytokine storm in COVID-19 pathogenesis [41].

Over the past two years, link between cholesterol metabolism and severity of COVID-19 was shown. As, it has also been demonstrated that COVID-19 disease is accompanied by dyslipidemia [4,5]. In particular, serum lipid levels decrease after the acute onset of COVID-19 and continue to decrease until the patient recovers [5]. Serum LDL levels have been found to be a predictor of poor disease prognosis of COVID-19 [6,42]. Patients with COVID-19 with a fatal outcome were characterized by irreversible decrease of LDL reduced by up to 60% compared with the level at hospitalization [42]. However, this study operated on a small cohort of patients with COVID-19 which included 21 patients with COVID-19 with concomitant chronic pathologies, among them only 4 cases of death. Aparisi and colleagues also demonstrated the association of low LDL serum level with 30-days all-cause mortality in patients with COVID-19 [43]. This study included representative sample number (654 patients, 149 cases of death), but expected the patient group was heterogeneous with apparent prevalence of diabetes mellitus, dyslipidemia, as well as other comorbidities such as chronic kidney disease, ischemic heart disease, which were maximally excluded in our study. Our data generally confirmed these findings: LDL level was decreased 7 days after admission to the ICU in non-survivors compared to survivors in both main and replication studies and at time admission in replication cohort.

Additionally, to lipid profile we analyzed cholesterol containing particles in plasma primarily LDLs and extracellular vesicles by alternative method based on immunoadsorption combined with DLS. Our previous study linked production of exomeres to cellular cholesterol synthesis [13]. We and others believe now that exomeres are responsible for essential part of plasma cholesterol [11,13]. Conjoint level of particles with hydrodynamic radius of 20 nm (exomers and LDLs), the same peak after immunoadsorption of exomeres using antibodies against CD9 and HSP90 (LDL particles remain after cleaning) and antibodies against apolipoproteins B100 (exomeres remain after cleaning) were analyzed. Relative LDL level determined using this approach generally repeated LDL profiles in the studied groups.

At the same time, we observed marked decrease in total proportion of exomeres and LDL particles in non-survivors when compared with both survivors and the control group even at the time of admission to the ICU in both main and replication studies. The role of conjoint fraction of exomeres and LDL particles as predictor of fatal outcome of COVID-19 was also supported via multivariable regression analysis adjusted for hypertension, obesity, age and sex. Various opinions exist concerning specificity of decreased LDL level and COVID19 outcome. Different points of view suggests that a significant decrease in LDL cholesterol level is also a feature of mortality during sepsis [44] or, alternatively, ICU patients receiving membrane oxygenation but not lethality [45]. According to our data conjoin fraction of exomeres and LDL particles may be more accurate than simple estimation of plasma LDL cholesterol concentration at the time of admission to the ICU as a relevant predictor of fatal outcome. However, the nature of the decrease of these classes of particles quantity is unknown. Previous study showed that exomeres are enriched in ecto-domain fragments of ACE2 receptor and thus can bind to SARS-CoV-2 S protein S1 subunit [46]. The exact role of exomeres in SARS-CoV-2 infection needs further study. At the same time the main result of our study allowed to suggest that overall LDL cholesterol assessed along with exomeres detected by DLS is more sensitive predictor of lethality during COVID19 infection compared to LDL level measured by standard technique.

## Conclusion

The current study confirmed that changes in the lipid profile against the back-ground of a cytokine storm may play a role in the pathogenesis of COVID-19. The severity of these changes may correlate with the outcome of the disease. The group of patients with severe COVID-19 with a poor outcome were characterized by the negative dynamics of HDL and LDL plasma levels during COVID-19. Decreased conjoint fraction of exomeres and LDL particles were identified as predictor of a fatal outcome of COVID-19. Further studies are needed to analyze if this observation could be referred to other critical illnesses or is specific to COVID-19 infection.

## Data Availability

All relevant data are within the manuscript and its Supporting Information files.

## Data Availability Statement

The original contributions presented in the study are included in the article/Supplementary Material, further inquiries can be directed to the corresponding author/s.

## Ethics Statement

The studies involving human participants were reviewed and approved by the Huizhou Central People’s Hospital. Written informed consent for participation was not required for this study in accordance with the national legislation and the institutional requirements. The study was approved by the Ethics Committee of the Pavlov First State Medical University of St. Petersburg (Russia).

## Author Contributions

T.U., S.P. – conceptualization, T.U. - writing—original draft preparation, S.P., V.M., S.L., A.T., P.S., Y.P.- writing—review and editing, T.U., A.B., V.M. - visualization, T.U., A.B., K.B., M.N., A.I., E.G., I.S. – sample collection, T.U., A.B., K.B., S.L., Z.K., N.L., M.N., A.I., E.G., I.S., I.V.E.C., Y.K. – investigation. All authors have read and agreed to the published version of the manuscript.

## References

1. Berekaa, M.M. Insights into the COVID-19 Pandemic: Origin, Pathogenesis, Diagnosis, and Therapeutic Interventions. Front. Biosci. (Elite Ed). 2021;13: 117–139. doi: 10.2741/874.

2. Montazersaheb S, Hosseiniyan Khatibi SM, Hejazi MS, Tarhriz V, Farjami A, Ghasemian Sorbeni F, Farahzadi R, Ghasemnejad T. COVID-19 Infection: An Overview on Cytokine Storm and Related Interventions. Virol. J. 2022;19: 92. doi: 10.1186/s12985-022-01814-1.

3. Alharthy A, Aletreby W, Faqihi F, Balhamar A, Alaklobi F, Alanezi K, Jaganathan P, Tamim H, Alqahtani SA, Karakitsos D, Memish ZA. Clinical Characteristics and Predictors of 28-Day Mortality in 352 Critically Ill Patients with COVID-19: A Retrospective Study. J. Epidemiol. Glob. Health 2021;11: 98–104. doi: 10.2991/jegh.k.200928.001.

4. Wei X, Zeng W, Su J, Wan H, Yu X, Cao X, Tan W, Wang H. Hypolipidemia Is Associated with the Severity of COVID-19. J. Clin. Lipidol. 2020;14: 297–304. doi: 10.1016/j.jacl.2020.04.008.

5. Sorokin AV, Karathanasis SK, Yang ZH, Freeman L, Kotani K, Remaley AT. COVID-19-Associated Dyslipidemia: Implications for Mechanism of Impaired Resolution and Novel Therapeutic Approaches. FASEB J. Off. Publ. Fed. Am. Soc. Exp. Biol. 2020;34: 9843–9853. doi: 10.1096/fj.202001451.

6. Hu X, Chen D, Wu L, He G, Ye W. Declined Serum High Density Lipoprotein Cholesterol Is As-sociated with the Severity of COVID-19 Infection. Clin. Chim. Acta. 2020;510: 105–110. doi: 10.1016/j.cca.2020.07.015.

7. Vlasov I, Panteleeva A, Usenko T, Nikolaev M, Izumchenko A, Gavrilova E, Shlyk I, Miroshnikova V, Shadrina M, Polushin Y, Pchelina S, Slonimsky P. Transcriptomic Profiles Reveal Downregulation of Low-Density Lipoprotein Particle Receptor Pathway Activity in Patients Surviving Severe COVID-19. Cells. 2021;10: 3495. doi: 10.3390/cells10123495.

8. Sirotkina OV, Ulitina AS, Kulabukhova DG, Nikolaev MA, Izyumchenko AD, Garaeva LA, Shlyk IV, Gavrilova EG, Polushin YuS, Pchelina SN. Correlation of laboratory markers of hemo-static system activation with concentration and size of plasma extracellular microparticles in patients with COVID-19. The Scientific Notes of the Pavlov University. 2022;29(1): 28–36. doi: 10.24884/1607-4181-2022-29-1-28-36. (In Russ.)

9. Krishnamachary B, Cook C, Spikes L, Chalise P, Dhillon NK. Extracellular vesicle-mediated endothelial apoptosis and EV-associated proteins correlate with COVID-19 disease severity. J. Extracell. Vesicles. 2021; 10(9): e12117. doi: 10.1002/jev2.12117.

10. Sun B, Tang N, Peluso MJ, Iyer NS, Torres L, Donatelli JL, Munter SE, Nixon CC, Rutishauser RL, Rodriguez-Barraquer I, Greenhouse B, Kelly JD, Martin JN, Deeks SG, Henrich TJ, Pulliam L. Characterization and Biomarker Analyses of Post-COVID-19 Complications and Neurological Manifestations. Cells. 2021;10: 386. doi: 10.3390/cells10020386.

11. Zhang H, Freitas D, Kim HS, Fabijanic K, Li Z, Chen H, Mark MT, Molina H, Martin AB, Bojmar L, Fang J, Rampersaud S, Hoshino A, Matei I, Kenific CM, Nakajima M, Mutvei AP, Sansone P, Buehring W, Wang H, Jimenez JP, Cohen-Gould L, Paknejad N, Brendel M, Manova-Todorova K, Magalhães A, Ferreira JA, Osório H, Silva AM, Massey A, Cubillos-Ruiz JR, Galletti G, Giannakakou P, Cuervo AM, Blenis J, Schwartz R, Brady MS, Peinado H, Bromberg J, Matsui H, Reis CA, Lyden D. Identification of Distinct Nanoparticles and Subsets of Extracellular Vesicles by Asymmetric Flow Field-Flow Fractionation. Nat. Cell Biol. 2018;20: 332–343. doi: 10.1038/s41556-018-0040-4.

12. Zhang Q, Higginbotham JN, Jeppesen DK, Yang YP, Li W, McKinley ET, Graves-Deal R, Ping J, Britain CM, Dorsett KA, Hartman CL, Ford DA, Allen RM, Vickers KC, Liu Q, Franklin JL, Bellis SL, Coffey RJ. Transfer of Functional Cargo in Exomeres. Cell Rep. 2019;27: 940-954.e6. doi: 10.1016/j.celrep.2019.01.009.

13. Landa S, Verlov N, Fedorova N, Filatov M, Pantina R, Burdakov V, Varfolomeeva E, Emanuel V. Extracellular Particles as Carriers of Cholesterol Not Associated with Lipoproteins. Membranes (Basel). 2022;12: 618. doi: 10.3390/membranes12060618.

14. Kanof ME, Smith PD, Zola H. Isolation of Whole Mononuclear Cells from Peripheral Blood and Cord Blood. Curr. Protoc. Immunol. 2001;Chapter 7: Unit 7.1. doi: 10.1002/0471142735.im0701s19.

15. Livak KJ, Schmittgen TD. Analysis of Relative Gene Expression Data Using Real-Time Quantitative PCR and the 2-ΔΔCT Method. Methods. 2001;25: 402–408. doi: 10.1006/meth.2001.1262.

16. Landa SB, Korabliov PV, Semenova EV, Filatov MV. Peculiarities of the Formation and Subsequent Removal of the Circulating Immune Complexes from the Bloodstream during the Process of Digestion. F1000Research. 2018;7: 618. doi: 10.12688/f1000research.14406.1.

17. Lebedev AD, Ivanova MA, Lomakin AV, Noskin VA. Heterodyne Quasi-Elastic Light-Scattering Instrument for Biomedical Diagnostics. Appl. Opt. 1997;36: 7518–7522. doi: 10.1364/ao.36.007518.

18. Ucciferri C, Caiazzo L, Di Nicola M, Borrelli P, Pontolillo M, Auricchio A, Vecchiet J, Falasca K. Parameters associated with diagnosis of COVID-19 in emergency department. Immunity, inflammation and disease. 2021;9(3): 851–861. doi: 10.1002/iid3.440.

19. Martín-Fernández M, Aller R, Heredia-Rodríguez M, Gómez-Sánchez E, Martínez-Paz P, Gonzalo-Benito H, Sánchez-de Prada L, Gorgojo Ó, Carnicero-Frutos I, Tamayo E, Tamayo-Velasco Á. Lipid Peroxidation as a Hallmark of Severity in COVID-19 Patients. Redox Biol. 2021;48: 102181. doi: 10.1016/j.redox.2021.102181.

20. Taban Q, Mumtaz PT, Masoodi KZ, Haq E, Ahmad SM. Scavenger Receptors in Host Defense: From Functional Aspects to Mode of Action. Cell Commun. Signal. 2022;20: 2. doi: 10.1186/s12964-021-00812-0.

21. Law AL, Ling Q, Hajjar KA, Futter CE, Greenwood J, Adamson P, Wavre-Shapton ST, Moss SE, Hayes MJ. Annexin A2 Regulates Phagocytosis of Photoreceptor Outer Segments in the Mouse Retina. Mol. Biol. Cell. 2009;20: 3896–3904. doi: 10.1091/mbc.e08-12-1204.

22. Zhang S, Yu M, Guo Q, Li R, Li G, Tan S, Li X, Wei Y, Wu M. Annexin A2 Binds to Endosomes and Negatively Regulates TLR4-Triggered Inflammatory Responses via the TRAM-TRIF Pathway. Sci. Rep. 2015;5: 15859. doi: 10.1038/srep15859.

23. Drücker P, Pejic M, Galla HJ, Gerke V. Lipid Segregation and Membrane Budding Induced by the Peripheral Membrane Binding Protein Annexin A2. J. Biol. Chem. 2013;288: 24764–24776. doi: 10.1074/jbc.M113.474023.

24. Maréchal L, Laviolette M, Rodrigue-Way A, Sow B, Brochu M, Caron V, Tremblay A. The CD36-PPARγ Pathway in Metabolic Disorders. Int. J. Mol. Sci. 2018;19: 1529. doi: 10.3390/ijms19051529.

25. AbdelMassih AF, Menshawey R, Ismail JH, Husseiny RJ, Husseiny YM, Yacoub S, Kamel A, Hozaien R, Yacoub E, Menshawey E, Abdelmalek A, Abouelazaem A, Elhatw A, Aboelmaaty A, Shahib A, Mansour A, Kamal A, Mohamed B, Atif B, Ghabreal B, Abdelmalak C, Ibrahim D, Elsaify E, Magdy F, Hanna FG, Hafez H, Dahir H, Merhom K, Ahmed M, Bishara M, Tawfik M, Youssef M, El Sharnouby M, Hamouda M, Ammar M, Ali N, Daniel N, El-Husseiny N, Abdelraouf N, Abdelhameed NK, Ahmed R, Othman R, Mohamadein R, Allam R, Elgendy R, Shebl R, Elsherbiney S, Fouad S, Emel S, Owais S, Hetta S, El-Saman S, Abdelalim S, Galal S, Asar Y, Osman Y, Khalaf Y, Aziz Y, Khafagy Y, Gamal N, Castaldi B. PPAR Agonists as Effective Adjuvants for COVID-19 Vaccines, by Modifying Immunogenetics: A Review of Literature. J. Genet. Eng. Biotechnol. 2021;19: 82. doi: 10.1186/s43141-021-00179-2.

26. Desterke C, Turhan AG, Bennaceur-Griscelli A, Griscelli F. PPARγ Cistrome Repression during Activation of Lung Monocyte-Macrophages in Severe COVID-19. iScience. 2020;23: 101611. doi: 10.1016/j.isci.2020.101611.

27. Hernandez-Quiles M, Broekema MF, Kalkhoven E. PPARgamma in Metabolism, Immunity, and Cancer: Unified and Diverse Mechanisms of Action. Front. Endocrinol. (Lausanne). 2021;12: 624112. doi: 10.3389/fendo.2021.624112.

28. Zhang H, Alford T, Liu S, Zhou D, Wang J. Influenza Virus Causes Lung Immunopathology through Down-Regulating PPARγ Activity in Macrophages. Front. Immunol. 2022;13: 958801. doi: 10.3389/fimmu.2022.958801.

29. Fantacuzzi M, Amoroso R, Ammazzalorso A. PPAR Ligands Induce Antiviral Effects Targeting Per-turbed Lipid Metabolism during SARS-CoV-2, HCV, and HCMV Infection. Biology (Basel). 2022;11: 114. doi: 10.3390/biology11010114.

30. Korobova ZR, Arsentieva NA, Liubimova NE, Dedkov VG, Gladkikh AS, Sharova AA, Chernykh EI, Kashchenko VA, Ratnikov VA, Gorelov VP, Stanevich OV, Kulikov AN, Pevtsov DE, Totolian AA. A Comparative Study of the Plasma Chemokine Profile in COVID-19 Patients Infected with Different SARS-CoV-2 Variants. Int. J. Mol. Sci. 2022;23(16): 9058. doi: 10.3390/ijms23169058.

31. Kudryavtsev IV, Arsentieva NA, Korobova ZR, Isakov DV, Rubinstein AA, Batsunov OK, Khamitova IV, Kuznetsova RN, Savin TV, Akisheva TV, Stanevich OV, Lebedeva AA, Vorobyov EA, Vorobyova SV, Kulikov AN, Sharapova MA, Pevtsov DE, Totolian AA. Heterogenous CD8+ T Cell Mat-uration and ‘Polarization’ in Acute and Convalescent COVID-19 Patients. Viruses. 2022;14(9): 1906. doi: 10.3390/v.14091906.

32. Monserrat J, Gómez-Lahoz A, Ortega MA, Sanz J, Muñoz B, Arévalo-Serrano J, Rodríguez JM, Gasalla JM, Gasulla Ó, Arranz A, Fortuny-Profitós J, Mazaira-Font FA, Teixidó Román M, Martínez-A C, Balomenos D, Asunsolo A, Álvarez-Mon M, On Behalf Of The Covid-Hupa Group. Role of Innate and Adaptive Cytokines in the Survival of COVID-19 Patients. International journal of molecular sciences. 2022;23(18): 10344. doi: 10.3390/ijms231810344.

33. Iwasaki A, Medzhitov R. Control of Adaptive Immunity by the Innate Immune System. Nat. Immunol. 2015;16: 343–353. doi: 10.1038/ni.3123.

34. Galani IE, Rovina N, Lampropoulou V, Triantafyllia V, Manioudaki M, Pavlos E, Koukaki E, Fragkou PC, Panou V, Rapti V, Koltsida O, Mentis A, Koulouris N, Tsiodras S, Koutsoukou A, Andreakos E. Untuned Antiviral Immunity in COVID-19 Revealed by Temporal Type I/III Interferon Patterns and Flu Comparison. Nat. Immunol. 2021;22: 32–40. doi: 10.1038/s41590-020-00840-x.

35. Del Valle DM, Kim-Schulze S, Huang HH, Beckmann ND, Nirenberg S, Wang B, Lavin Y, Swartz TH, Madduri D, Stock A, Marron TU, Xie H, Patel M, Tuballes K, Van Oekelen O, Rahman A, Kovatch P, Aberg JA, Schadt E, Jagannath S, Mazumdar M, Charney AW, Firpo-Betancourt A, Mendu DR, Jhang J, Reich D, Sigel K, Cordon-Cardo C, Feldmann M, Parekh S, Merad M, Gnjatic S. An Inflammatory Cytokine Signature Predicts COVID-19 Severity and Survival. Nat. Med. 2020;26: 1636–1643. doi: 10.1038/s41591-020-1051-9.

36. Lucas C, Wong P, Klein J, Castro TBR, Silva J, Sundaram M, Ellingson MK, Mao T, Oh JE, Israelow B, Takahashi T, Tokuyama M, Lu P, Venkataraman A, Park A, Mohanty S, Wang H, Wyllie AL, Vogels CBF, Earnest R, Lapidus S, Ott IM, Moore AJ, Muenker MC, Fournier JB, Campbell M, Odio CD, Casanovas-Massana A; Yale IMPACT Team, Herbst R, Shaw AC, Medzhitov R, Schulz WL, Grubaugh ND, Dela Cruz C, Farhadian S, Ko AI, Omer SB, Iwasaki A. Longitudinal Analyses Reveal Immunological Misfiring in Severe COVID-19. Nature. 2020;584: 463–469. doi: 10.1038/s41586-020-2588-y.

37. Grant RA, Morales-Nebreda L, Markov NS, Swaminathan S, Querrey M, Guzman ER, Abbott DA, Donnelly HK, Donayre A, Goldberg IA, Klug ZM, Borkowski N, Lu Z, Kihshen H, Politanska Y, Sichizya L, Kang M, Shilatifard A, Qi C, Lomasney JW, Argento AC, Kruser JM, Malsin ES, Pickens CO, Smith SB, Walter JM, Pawlowski AE, Schneider D, Nannapaneni P, Abdala-Valencia H, Bharat A, Gottardi CJ, Budinger GRS, Misharin AV, Singer BD, Wunderink RG; NU SCRIPT Study Investigators. Circuits between Infected Macrophages and T Cells in SARS-CoV-2 Pneumonia. Nature. 2021;590: 635–641. doi: 10.1038/s41586-020-03148-w.

38. Arsentieva NA, Liubimova NE, Batsunov OK, Korobova ZR, Stanevich OV, Lebedeva AA, Vorobyov EA, Vorobyova SV, Kulikov AN, Lioznov DA, Sharapova MA, Pevtcov DE, Totolian AA. Plasma Cytokines in Patients with COVID-19 during Acute Phase of the Disease and Following Complete Recovery. Med. Immunol (Russia). 2021;23: 311–326 (in Russ.) doi: 10.15789/1563-0625-PCI-2312. (in Russ.)

39. Vu CN, Ruiz-Esponda R, Yang E, Chang E, Gillard B, Pownall HJ, Hoogeveen RC, Coraza I, Balasubramanyam A. Altered Relationship of Plasma Triglycerides to HDL Cholesterol in Patients with HIV/HAART-Associated Dyslipidemia: Further Evidence for a Unique Form of Metabolic Syndrome in HIV Patients. Metabolism. 2013;62: 1014–1020. doi:10.1016/j.metabol.2013.01.020.

40. Dias SSG, Soares VC, Ferreira AC, Sacramento CQ, Fintelman-Rodrigues N, Temerozo JR, Teixeira L, Nunes da Silva MA, Barreto E, Mattos M, de Freitas CS, Azevedo-Quintanilha IG, Manso PPA, Miranda MD, Siqueira MM, Hottz ED, Pão CRR, Bou-Habib DC, Barreto-Vieira DF, Bozza FA, Souza TML, Bozza PT. Lipid droplets fuel SARS-CoV-2 rep-lication and production of inflammatory mediators. PLoS pathogens. 2022;16(12): e1009127. doi: 10.1371/journal.ppat.1009127.

41. Caterino M, Gelzo M, Sol S, Fedele R, Annunziata A, Calabrese C, Fiorentino G, D’Abbraccio M, Dell’Isola C, Fusco FM, Parrella R, Fabbrocini G, Gentile I, Andolfo I, Capasso M, Costanzo M, Daniele A, Marchese E, Polito R, Russo R, Missero C, Ruoppolo M, Castaldo G. Dysregulation of lipid metabolism and pathological inflammation in pa-tients with COVID-19. Scientific reports. 2021;11(1): 2941. doi: 10.1038/s41598-021-82426-7.

42. Fan J, Wang H, Ye G, Cao X, Xu X, Tan W, Zhang Y. Letter to the Editor: Low-Density Lipoprotein Is a Potential Predictor of Poor Prognosis in Patients with Coronavirus Disease 2019. Metabolism. 2020;107: 154243. doi: 10.1016/j.metabol.2020.154243.

43. Aparisi Á, Iglesias-Echeverría C, Ybarra-Falcón C, Cusácovich I, Uribarri A, García-Gómez M, Ladrón R, Fuertes R, Candela J, Tobar J, Hinojosa W, Dueñas C, González R, Nogales L, Calvo D, Carrasco-Moraleja M, San Román JA, Amat-Santos IJ, Andaluz-Ojeda D. Low-Density Lipoprotein Cholesterol Levels Are Associated with Poor Clinical Outcomes in COVID-19. Nutr. Metab. Cardiovasc. Dis. 2021;31: 2619–2627. doi: 10.1016/j.numecd.2021.06.016.

44. Tanaka, S.; De Tymowski, C.; Stern, J.; Bouzid, D.; Zappella, N.; Snauwaert, A.; Robert, T.; Lortat-Jacob, B.; Tran-Dinh, A.; Augustin, P.; et al. Relationship between liver dysfunction, lipoprotein concentration and mortality during sepsis. PloS one, 2022, 17(8), e0272352, doi:10.1371/journal.pone.0272352.

45. Tanaka S, De Tymowski C, Zappella N, Snauwaert A, Robert T, Lortat-Jacob B, Castier Y, Tran-Dinh A, Tashk P, Bouzid D, Para M, Pellenc Q, Atchade E, Meilhac O & Montravers P. Lipoprotein concentration in patients requiring extracorporeal membrane oxygenation. Scientific reports. 2021;11(1): 17225. doi: 10.1038/s41598-021-96728-3.

46. Zhang Q, Jeppesen DK, Higginbotham JN, Franklin JL, Crowe JE Jr, Coffey RJ. Angioten-sin-Converting Enzyme 2-Containing Small Extracellular Vesicles and Exomeres Bind the Severe Acute Respiratory Syndrome Coronavirus 2 Spike Protein. Gastroenterology. 2021;160: 958-961.e3. doi: 10.1053/j.gastro.2020.09.042.

